# Identifying an oculomotor phenotype for adolescent depression with an interleaved pro- and anti-saccade task

**DOI:** 10.64898/2026.02.20.26346728

**Authors:** Blake K. Noyes, Linda Booij, Heidi C. Riek, Brian C. Coe, Don C. Brien, Sarosh Khalid-Khan, Douglas P. Munoz

**Affiliations:** Centre for Neuroscience Studies, Queen’s University, Kingston, Ontario, Canada; Department of Psychiatry, McGill University, Montreal, Quebec, Canada; Eating Disorders Continuum & Douglas Research Centre, Montreal, Quebec, Canada; Department of Psychiatry, Queen’s University, Kingston, Ontario, Canada; Biomedical and Molecular Sciences, Queen’s University, Kingston, Ontario, Canada

**Author notes:** **Corresponding author:** Blake K. Noyes.

**Keywords:** mood disorders, eye-tracking, fixation, saccade, pupil

## Abstract

Numerous studies have shown that adults with depression have distinct oculomotor alterations during saccade tasks, but whether similar alterations occur in adolescents is largely unknown. The purpose of this study was to test if eye-tracking during a structured saccade task could distinguish a group of adolescents with depression from healthy controls. We hypothesized that, due to overlapping circuitry between depression pathology and the oculomotor system, adolescents with depression would show alterations in fixation, saccade, and pupil behaviour. 51 adolescents with depression and 66 age-matched healthy controls completed the Interleaved Pro- and Anti-Saccade Task (IPAST) and several self-reported questionnaires for psychiatric symptoms. Oculomotor outcomes included fixation acquisition, fixation breaks, correct rate, saccadic reaction time, rate of correct express-latency pro-saccades, rate of express- and regular-latency anti-saccade errors, baseline pupil size, as well as pupil constriction and dilation sizes following task instruction. In comparison to healthy controls, adolescents with depression displayed impairments acquiring fixation (*p*<.001), made more fixation breaks in pro- (*p*=.023) and anti-saccade trials *(p*=.005), more anti-saccade errors (*p*=.013), more express-latency saccades overall (*p*s=.016), had a smaller pupil constriction in pro-saccade trials (*p*=.047) and had a smaller pupil dilation in pro- (*p*=.011) and anti-saccade trials (*p*=.041). No differences were found for saccadic reaction time, rate of correct pro-saccades, rate of regular-latency anti-saccade errors, pupil constriction size during anti-saccade trials, or baseline pupil size. Patients had psychiatric comorbidities and were using psychotropic medication. While this reflected clinical reality, these factors may have influenced oculomotor behaviour. Adolescents with depression had altered fixation, saccade, and pupil behaviour during IPAST. Given that many cases of adolescent depression remain undetected, accessible and objective screening approaches are highly needed. This oculomotor phenotype may be used in the development of such a screening tool to detect those at risk.

## INTRODUCTION

Approximately one in five adolescents worldwide have depressive symptoms.^1^ These symptoms typically have an onset in mid- to late adolescence, and even a perceived “low” number of symptoms can have a severe impact on wellbeing and psychosocial functioning.^2^ Additionally, while depression itself is highly heterogenous, it is often accompanied by comorbid conditions such as anxiety or substance use disorders, further complicating diagnosis and clinical decision-making.^3^ Despite it being well known that early identification and treatment of mental health concerns is protective against worsening progression^4,5^, the majority of adolescents requiring care for mental health conditions such as depression go unrecognized or untreated.^6^ In addition to systemic barriers such as limitations to health care access, personal factors such as concerns about symptom disclosure, self-stigma, nervousness and ambivalence around seeking help can hinder timely diagnosis and treatment.^7^ Thus, there is a need for quick, objective, non-invasive, and cost-effective tools to identify youth at risk.

One possible approach to meet this need is to investigate the oculomotor behaviour of adolescents with depression. Oculomotor behaviour such as saccades (rapid eye movements from one location to another), visual fixation (maintaining gaze on a particular location), and pupil responses (constriction and dilation) have been studied extensively in healthy individuals. Across different eye-tracking tasks, normative oculomotor behaviour has been well-established and can be linked back to specific brain areas and neural circuits.^8^ What makes studying oculomotor behaviour relevant for the field of psychiatry is the substantial overlap in the established neurocircuitry involved in oculomotor control and the neurocircuitry altered in various psychiatric conditions.^9^ This overlap, coupled with the accessibility of video-based eye-tracking as a research tool, has made this a promising method to study psychiatric conditions.^10–12^

While various eye-tracking paradigms have been developed, eye-tracking during a pro- and anti-saccade task may be particularly advantageous, as this task probes sensory, motor, and cognitive components of neural circuitry which are commonly impacted in neurological and psychiatric conditions. A typical pro-saccade trial involves fixating in the center of a screen and then looking at a peripheral stimulus as soon as it appears. Pro-saccades are easy to execute, usually elucidating a quick, automatic response, which probes the integrity of the visual and motor components of the oculomotor circuit.^13,14^ In contrast, anti-saccades require the participant to look away from the peripheral stimulus. Like pro-saccade trials, they begin with a central fixation, but when the stimulus appears, the participant must look to the opposite side. This requires the participant to inhibit the automatic response to look towards the stimulus and then generate a voluntary saccade in the opposite direction, without a visual stimulus. The anti-saccade task evaluates the integrity of the circuitry required for coordination of voluntary movement, as well as cognitive control.^13,14^

Given that different psychiatric conditions may correlate with differential patterns of brain structure, function, and connectivity, it is reasonable to expect that each condition could be associated with a specific oculomotor phenotype. In depression, neuroimaging studies have found structural, functional and network connectivity alterations.^15^ This includes hypoactivity within the central executive network (CEN; also known as the frontoparietal network), which is responsible for executive functioning.^15^ Clinically, this hypoactivity is related to impairments in motivation, attention, and cognitive performance.^16^ However, this network is also implicated in oculomotor behaviour, especially during cognitive tasks. The dorsolateral prefrontal cortex (dlPFC) and frontal eye fields (FEF), areas within the CEN, are crucial for correct anti-saccade performance^17,18^, as well as pupillary response during pro- and anti-saccade tasks.^19,20^

Several studies have investigated pro- and/or anti-saccade behaviour between adults (18-60) with depression and healthy controls.^21–30^ The most frequently investigated oculomotor outcomes across these studies are correct rate (saccades made in the correct direction) and saccadic reaction time (SRT), which measures the time from stimulus appearance to the onset of the first saccade. Typically, depressed individuals have been found to have similar correct rate for pro-saccade trials and a decrease in correct rate for anti-saccade trials in comparison to healthy controls, whereas mixed results have been reported for SRT. Although these studies show that adults with depression have altered oculomotor behaviour during saccade tasks, it is unclear whether similar alterations occur in adolescents with depression. No pro-saccade data from adolescents with depression have been reported, and only a few studies have investigated anti-saccade performance^31,32^. However, these studies utilized a reward saccade task with monetary incentives, making it difficult to generalize these results to basic anti-saccade performance.

The aim of the present study is to investigate if eye-tracking during the Interleaved Pro- and Anti-Saccade Task (IPAST)^33^ could distinguish a group of adolescents with depression from healthy controls. While previous studies utilizing video-based eye tracking have primarily reported saccade parameters, we also investigated fixation control and pupil response. These have been found to be altered in other cognitively-impaired populations^11,34^, and these data would help build a fuller picture of depression’s oculomotor phenotype. We hypothesized that adolescents with depression will have increased anti-saccade error rates compared to healthy controls due to poor cognitive control. As an exploratory analysis, we also assessed level of fixation acquisition, rate of fixation breaks (indicative of task readiness/focus), and pupillary responses, all of which we expect to be altered in depression. Finally, we investigated whether there are associations between scores on clinical assessments and oculomotor behaviour. Should saccade, fixation, and/or pupil alterations be evident in adolescence, they may contribute to the future development of a new early-identification tool for this vulnerable population.

## METHOD

The study was approved by the Queen’s University Health Sciences Research Ethics Board (HSREB #6040314, formerly #6005163). Data collection occurred between April 2021 and January 2025. Adult participants and parents or guardians of participants under the age of 18 provided consent prior to commencing the study. Children provided assent.

### Participants

All participants were between the ages of 12-19 years, fluent in the English language, had normal or corrected-to-normal vision, and no neurological conditions (e.g., seizures). No participants in this study had previously completed this task or any other task in our laboratory.

#### Clinical Group

Adolescent patients with depression (DEP) were recruited from an outpatient mental health program at Kingston Health Sciences Centre. A primary diagnosis of a depressive disorder according to the Diagnostic and Statistical Manual of Mental Disorders, Fifth Edition (DSM-5) was required. Diagnoses were confirmed by an adolescent psychiatrist (SKK). Patients were not eligible to participate if they had comorbid psychosis, were engaged in extreme substance use, or had high levels of active suicidal ideation as confirmed by SKK.

#### Control Group

Healthy controls (HC) were recruited from the local community. Controls were required to have no past or present mental health disorders, and no past or present psychotropic medication usage. To confirm this, prospective controls were administered the MINI International Neuropsychiatric Interview (MINI; 18+)^35^ or MINI International Neuropsychiatric Interview for Children and Adolescents (MINI-KID; 12-17)^36^ by a trained experimenter (BKN). Scoring was verified by a clinical psychologist (LB) when required.

### Equipment and Materials

#### Eye-tracking

Monocular eye movements and pupil size were recorded in a dark room with an EyeLink 1000 Plus eye-tracker (SR Research, Ottawa) at a sampling rate of 500Hz. Participants were seated 60cm away from a 17-inch 1280×1024-pixel resolution LCD computer monitor with their head stabilized in a chin and forehead rest while gaze position was continuously tracked.

#### Clinical Assessments

Participants in the depression group and screened controls completed a set of standardized questionnaires evaluating a spectrum of psychiatric symptoms: Barratt Impulsiveness Scale (BIS-30; 30 items; higher scores indicate greater impulsivity)^37^, Borderline Symptom List (BSL-23; 23 items; higher scores indicate more symptoms of borderline personality disorder)^38^, Difficulties in Emotion Regulation Scale (DERS-36; 36 items; higher scores indicate decreased emotional regulation)^39^, Eating Disorder Examination Questionnaire (EDE-Q; 36 items; higher scores indicate more symptoms of eating disorders)^40^, Generalized Anxiety Disorder Scale (GAD-7; 7 items; higher scores indicate greater anxiety)^41^, Patient Health Questionnaire (PHQ-9; 9 items; higher scores indicate greater depression)^42^, Suicide Behaviors Questionnaire (SBQ-R; 4 items; higher scores greater risk of suicide)^43^. Participants were given the option of completing these at the eye-tracking session or online before/after, excluding the SBQ-R, which was always completed in person due to its sensitive nature.

### Task

IPAST consisted of two blocks of 120 trials, each of which were broken into four epochs (Fig. 1). Each trial began with a blank black screen for 1000ms (intertrial interval; ITI epoch). Then, a green or red central fixation point (0.5° diameter, 44 cd/m^2^) appeared for 1000ms (FIX epoch). The colour of the fixation point indicated whether it was a pro-saccade (green) or anti-saccade (red) trial. This was followed by the disappearance of the fixation point for 200ms (GAP epoch), until a gray peripheral stimulus (0.5° diameter, 62 cd/m^2^) appeared 10° to the left or right for 1000ms (STIM epoch). Participants were required to look at the stimulus (pro-saccade instruction) or in the opposite direction (anti-saccade instruction). Trial type (pro-/anti-saccade) and stimulus location (left/right) were pseudo-randomly interleaved for both blocks.

**FIGURE 1.**
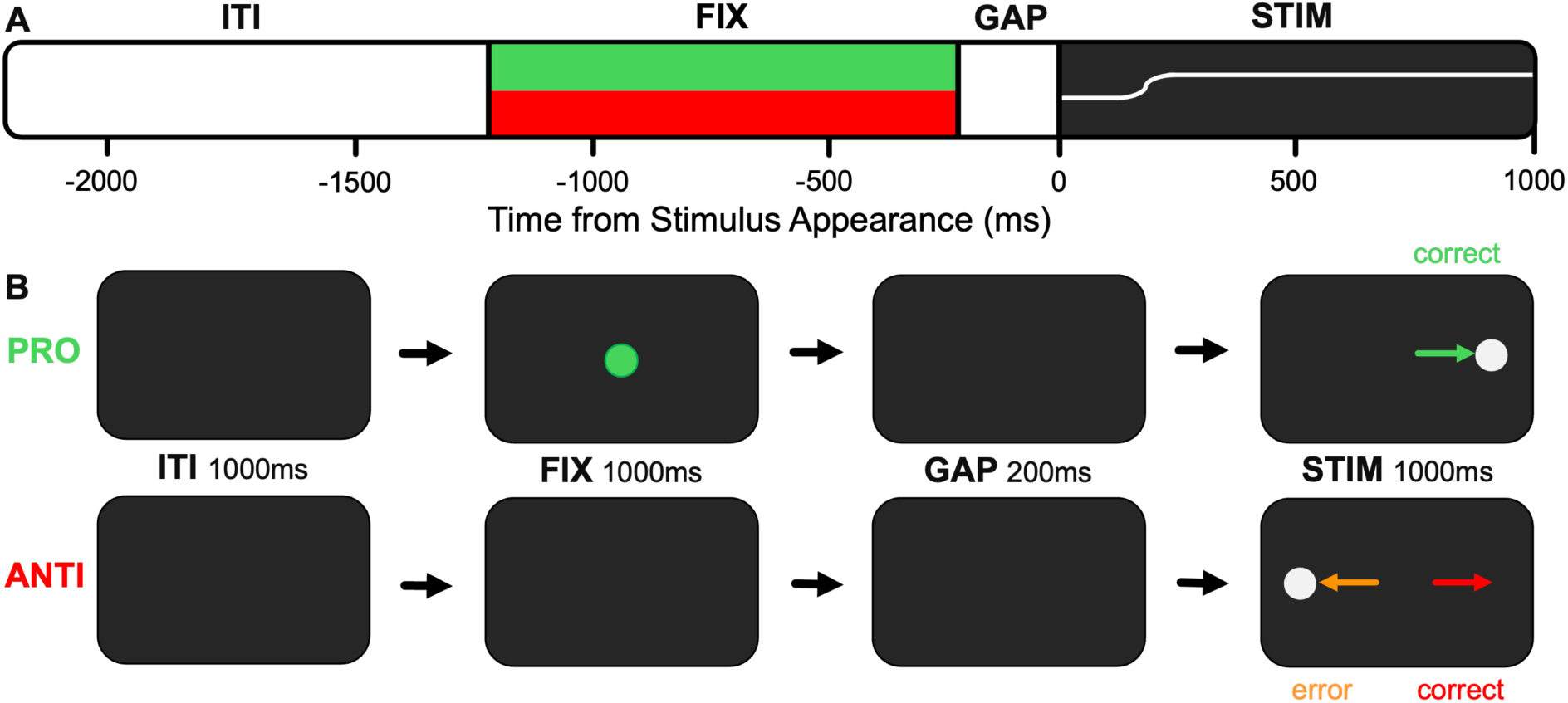
A. Timing of the Interleaved Pro- and Anti-Saccade Task (IPAST) epochs. B. Spatial representation of the task epochs (central fixation point and stimulus size not to scale). ITI=intertrial interval epoch, FIX=fixation epoch, GAP=gap epoch, STIM=stimulus/response epoch. PRO=pro­ saccade trial, ANTI=anti-saccade trial.

### Procedure

Prospective control participants completed the MINI or MINI-KID over Zoom prior to being invited to participate in the study. Eligible controls and patients were sent an online link to the study questionnaires 24-48 hours prior to the experimental session and were asked to complete them prior to attending. These questionnaires took 15-30 minutes to complete.

After arriving at the in-person session, the experimenter began with the eye-tracking portion of the study. Once the participant understood the task, they sat in front of the eye-tracker and adjusted the seat height to their comfort. A nine-point array was used to calibrate and validate eye position, and then IPAST was initiated (∼20 minutes). Additional calibrations were completed in response to movement or participant breaks as needed. Then, participants completed the SBQ-R (1 min) and any additional questionnaires not completed online. Participants were provided with a $50 gift card to a major retailer for their time.

### Data Analysis

#### Oculomotor Analysis

All oculomotor outcomes, including fixation, saccade, and pupil behaviour, were preprocessed using the laboratory’s standardized pipeline.^33,44,45^ Group differences in oculomotor behaviour were analyzed using three separate Multivariate Analysis of Variance (MANOVA) tests for fixation, saccade, and pupil outcomes, respectively.

Fixation outcomes included *rate of fixation acquisition*, and *rate of fixation breaks* for pro- and anti-saccade trials. Fixation acquisition was defined as the rate of trials where gaze was fixated in the center of the screen before the fixation point appeared, and was measured at the end of the ITI epoch (–1200ms from stimulus appearance).^34^ As it was assessed prior to fixation point appearance, this outcome was not separated by trial type. Fixation breaks were defined as rate of trials where the participant looked away from the central fixation point (saccade >2° amplitude) and did not return before the FIX epoch ended.^44^

Saccade outcomes included *correct rate* for pro- and anti-saccade trials, *rate of correct express-latency saccades* for pro-saccade trials, *rate of express-latency errors* for anti-saccade trials, *rate of regular-latency errors* for anti-saccade trials, and *saccadic reaction time* (SRT; ms) for pro- and anti-saccade trials. We separated anti-saccade errors into express- and regular-latency as they measure different underlying neural processes.^13^ Express-latency saccades are saccades 90-139ms after stimulus onset and represent the earliest visually guided saccades, which are made prior to top-down control signals being received. Regular-latency saccades are those made 140-800ms after STIM onset and can be influenced by top-down control. Therefore, express-latency saccades impair performance for anti-saccade trials, but not pro-saccade trials. All saccade analysis used the first saccade after stimulus onset, and only included saccades made during the viable reaction time window (90-800ms from stimulus onset).^33^ This eliminated anticipatory saccades (guessing behaviour; -110-89 from stimulus onset) as well as late saccades (indicative of inattention; >800ms).

Pupil outcomes included *baseline pupil size, constriction size* during pro- and anti-saccade trials, and *dilation size* during pro- and anti-saccade trials (see Supplementary Methods and Supplementary Fig. 1). During IPAST, a consistent pattern of pupil behaviour emerges: after fixation point appearance, the pupil constricts, quickly followed by a dilation response.^46^ Baseline pupil size was defined as the average pupil size (pixels) 200-300ms after fixation onset. We define pupil constriction as the difference between the pupil size at peak constriction and the maximum size prior to constriction. Pupil dilation was defined as the difference between the pupil size at peak dilation (end of the GAP epoch) and size at peak constriction. To remove the influence of individual/group differences in baseline pupil size on constriction and dilation sizes, the values (pixels) were first normalized to baseline pupil size, thus converting the data into relative percentage change.

#### Correlations

We assessed the relation between the fifteen oculomotor outcomes and measures of impulsivity (BIS-30), borderline traits (BSL-23), emotional regulation (DERS-36), eating disorder symptoms (EDE-Q), anxiety symptoms (GAD-7), depression symptoms (PHQ-9), and suicidality (SBQ-R). Pearson correlations were performed within the DEP group only (n=51) as well as for the entire sample (N=117).

#### Medication Effects

Finally, we examined the effects of medication on oculomotor behaviours within the DEP group. Medications were grouped into three classes (antidepressants, antipsychotics, and stimulants) and coded as ON/OFF. Similar to above, these were analyzed using three separate MANOVAs for fixation, saccade, and pupil outcomes with medication classes as fixed factors.

## RESULTS

### Participant Demographics

Twenty prospective controls were excluded for meeting MINI/MINI-KID criteria for a past or present psychiatric condition (n=18), meeting clinical levels of depression on the online questionnaires (PHQ score=22; n=1), and disclosure of a neurological/psychiatric condition following data collection (n=1). The final sample included 51 adolescents with depression (DEP; 40 female; mean age=15.88, *SD*=1.46) and 66 healthy controls (HC; 51 female; mean age=16.01, *SD*=2.20). HC and DEP did not differ on total age, female age, male age, or sex distribution (all *p*>.725). Adolescents with DEP scored higher on all clinical assessments (*p*s<.001; see Table 1) than the control group. DEP diagnoses included major depressive disorder (n=24; 47.06%), persistent depressive disorder (n=4; 7.84%), MDD and PDD (n=2; 3.92%) bipolar disorder in depressive phase (n=7; 13.73%), other specified depressive disorder (n=13; 25.49%), and unspecified depressive disorder (n=1; 1.96%). The majority of patients had at least one comorbidity, including anxiety disorders (n=28; 54.90%), developmental disorders (n=38; 74.51%), disruptive disorders (n=1; 1.96%), eating disorders (n=1; 1.96%), obsessive compulsive disorder (n=2; 3.92%), posttraumatic stress disorder (n=2; 3.92%), substance use disorders (n=1; 1.96%), and traits of borderline personality disorder (n=7; 13.73%),.

**Table 1.** Demographic and clinical characteristics of the study groups. HC=healthy controls, DEP=depression group, BIS=Barratt Impulsiveness Scale, BSL=Borderline Symptom List, DERS=Difficulties in Emotion Regulation Scale, EDE-Q=Eating Disorder Examination Questionnaire, GAD=Generalized Anxiety Disorder Scale, PHQ=Patient Health Questionnaire, SBQ-R=Suicide Behaviors Questionnaire Revised. **Significant at the p<.01 level. ^†^Two adolescents in the DEP group are missing SBQ-R scores due to incomplete questionnaires.

|  | HC (n=66) | DEP (n=51) |  |  |  |  |
| --- | --- | --- | --- | --- | --- | --- |
| | *M (SD)* | *M (SD)* | *F* | *df* | *p* | $\eta^2p$ |
| **Demographics** |  |  |  |  |  |  |
| Group age | 16.01(2.20) | 15.88(1.46) | .123 | 1, 115 | .727 | .001 |
| Female age | 16.00(2.33) | 15.85(1.35) | .125 | 1, 89 | .725 | .001 |
| Male age | 16.04(1.74) | 16.00(1.87) | .004 | 1, 24 | .951 | .000 |
| Percent female | 77.27% | 78.43% | .022 | 1, 115 | .882 | .000 |
| **Clinical Assessments**<br>**(possible score range)** |  |  |  |  |  |  |
| BIS-30 (30-120) | 59.24(9.54) | 74.86(10.11) | 73.172 | 1, 115 | <.001** | .389 |
| BSL-23 (0-4) | 0.15(0.17) | 1.55(0.82) | 182.065 | 1, 115 | <.001** | .613 |
| DERS-36 (36-180) | 66.77(15.77) | 119.71(26.07) | 184.801 | 1, 115 | <.001** | .616 |
| EDE-Q (0-6) | 0.66(0.93) | 1.94(1.59) | 29.443 | 1, 115 | <.001** | .204 |
| GAD-7 (0-21) | 1.71(2.68) | 12.14(4.87) | 217.454 | 1, 115 | <.001** | .654 |
| PHQ-9 (0-27) | 2.09(2.69) | 13.92(5.65) | 224.013 | 1, 115 | <.001** | .661 |
| SBQ-R<sup>†</sup> (3-18) | 3.48(1.19) | 10.29(4.47) | 139.913 | 1, 113 | <.001** | .553 |

### IPAST Behaviour

All group level comparison data are detailed in Supplementary Table 1.

#### Fixation Behaviour

Figure 2A displays the cumulative percentage of fixation acquisition over time. HC had a higher percentage of fixation acquisition (*F*(1,115)=16.136, *p*<.001, *η²p*=.123) than DEP at fixation appearance (-1200ms). DEP had a higher rate of fixation breaks during both pro- (Fig. 2B; *F*(1,115)=5.341, *p*=.023, *η²p*=.044) and anti-saccade trials (Fig. 2C; *F*(1,115)=8.384, *p*=.005, *η²p*=.068).

**FIGURE 2.**
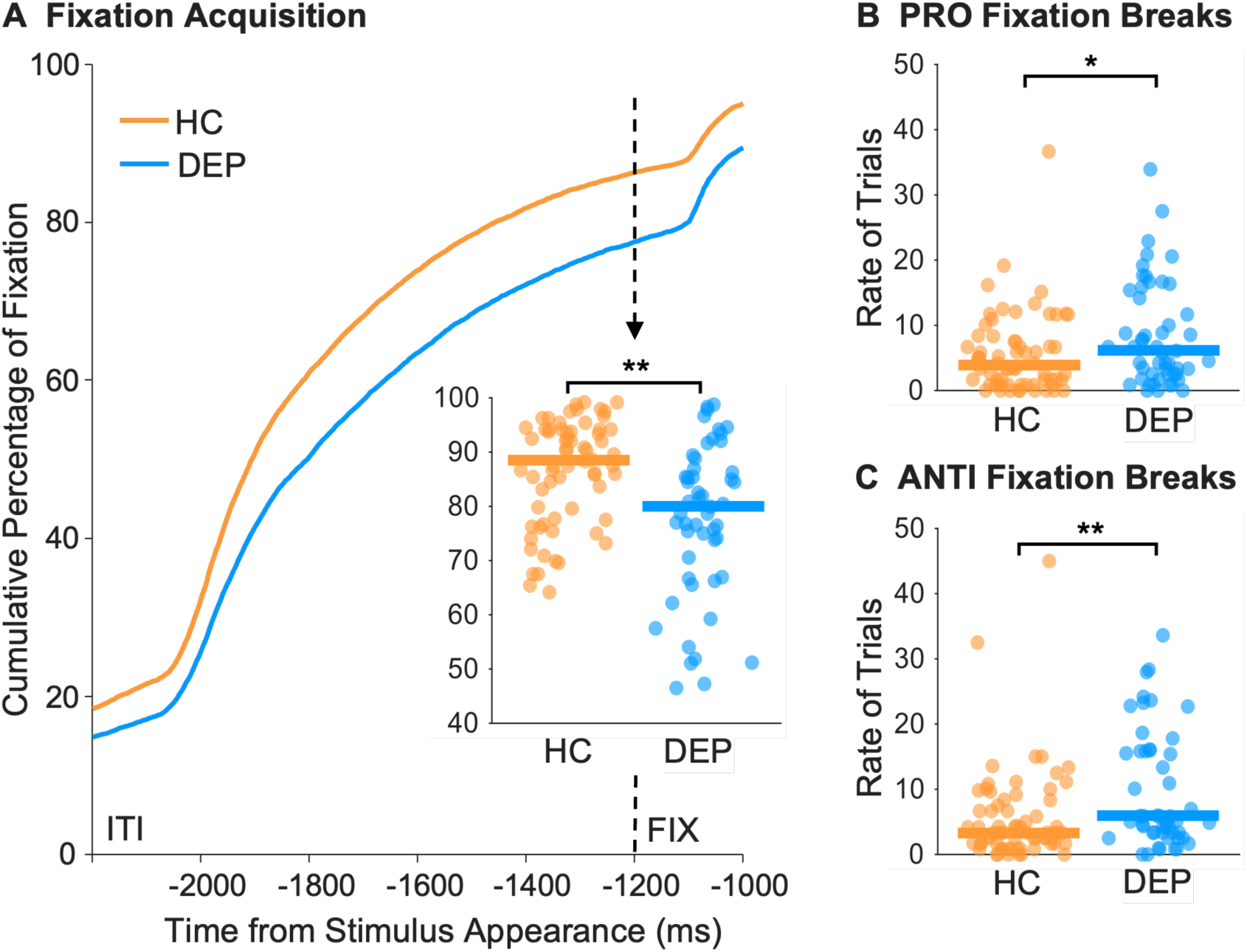
A. Cumulative percentage of fixation acquisition over time during the ITI. Inset bubble plot displays the difference at FIX onset (-1200ms). **B, C.** Rate of fixation breaks during pro- and anti-saccade trials, respectively for individual subjects. Horizontal bars represents the median group value. HC=healthy control, DEP=depression. *Significant at p<.05 level. ** Significant at p<.01 level.

#### Saccade Behaviour

Figure 3A shows the cumulative percentage of correct and incorrect saccades made over time for pro- and anti-saccade trials. DEP made more express-latency saccades in pro-saccade trials (*F*(1,115)=6.310, *p*=.016, *η²p*=.052) and express-latency errors during anti-saccade trials (Fig. 3B; *F*(1,115)=5.930, *p*=.016, *η²p*=.049). DEP made more anti-saccade errors overall (*F*(1,115)=6.378, *p*=.013, *η²p*=.053). HC and DEP did not differ on SRT for pro- or anti-saccade trials, correct rate for pro-saccade trials, or rate of regular-latency errors in anti-saccade trials (Fig. 3C).

**FIGURE 3.**
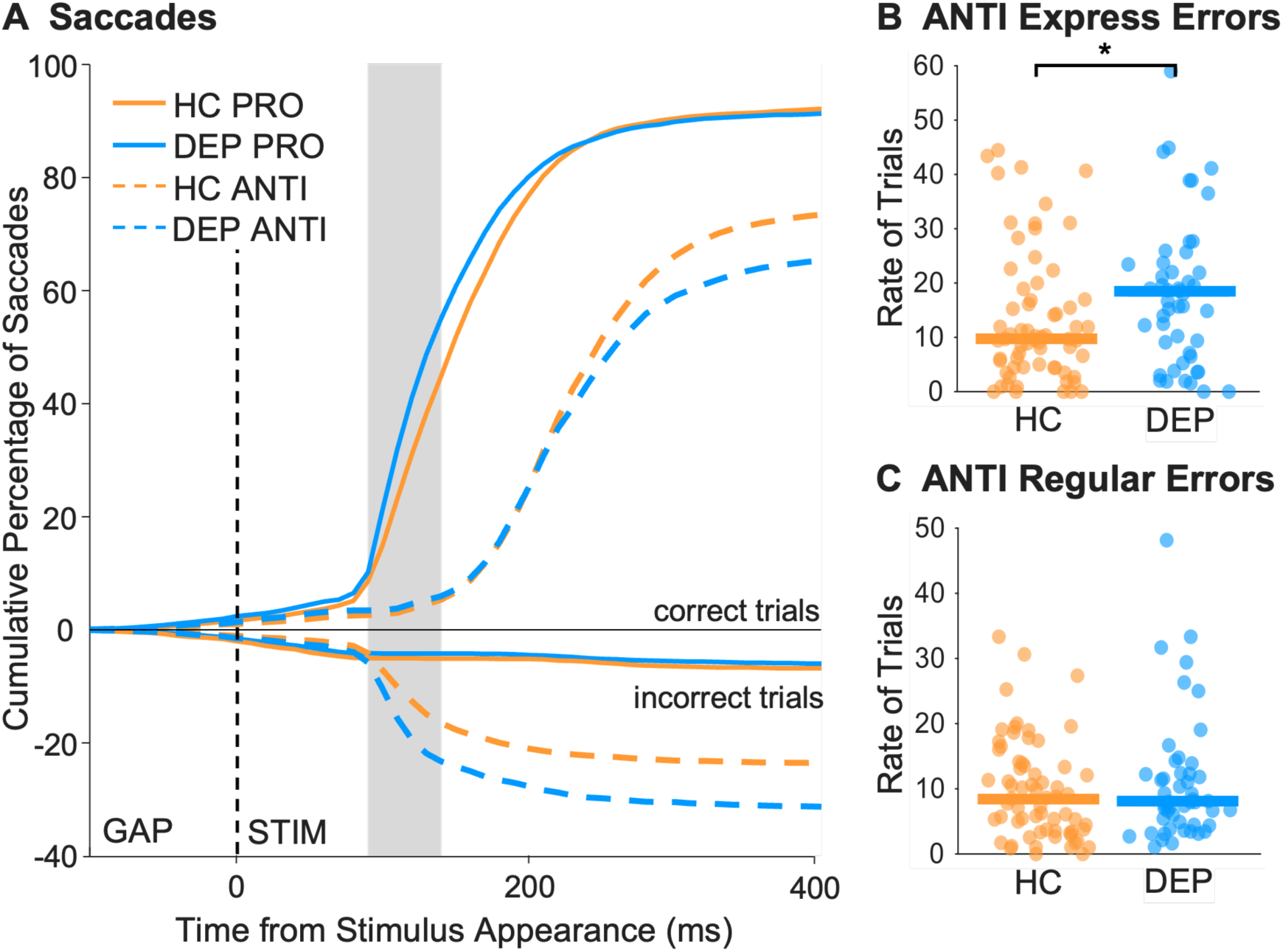
A. The cumulative distribution of correct and error saccadic reaction times made during pro- and anti-saccade trials following stimulus appearance. Positive and negative values on the y-axis indicate correct and incorrect direction saccades, respectively. The grey bar indicates the express-latency saccade window (90-140ms). **B.** Rate of express-latency errors in anti-saccade trials. C. Rate of regular-latency errors in anti-saccade trials. Horizontal lines represents the median group value. *Significant at p<.05 level.

#### Pupil Behaviour

Overall, the pattern for pupil behaviour was similar between groups, with an initial constriction after fixation appearance followed quickly by a dilation response before stimulus appearance (Fig. 4A). For pro-saccade trials, DEP pupil constriction size (*F*(1,115)=4.039, *p*=.047, *η²p*=.034) and dilation size (Fig. 4B; *F*(1,115)=6.768, *p*=.011, *η²p*=.056) were smaller than HC. For anti-saccade trials, only dilation size was smaller (Fig. 4C; *F*(1,115)=4.251, *p*=.041, *η²p*=.036).

**FIGURE 4.**
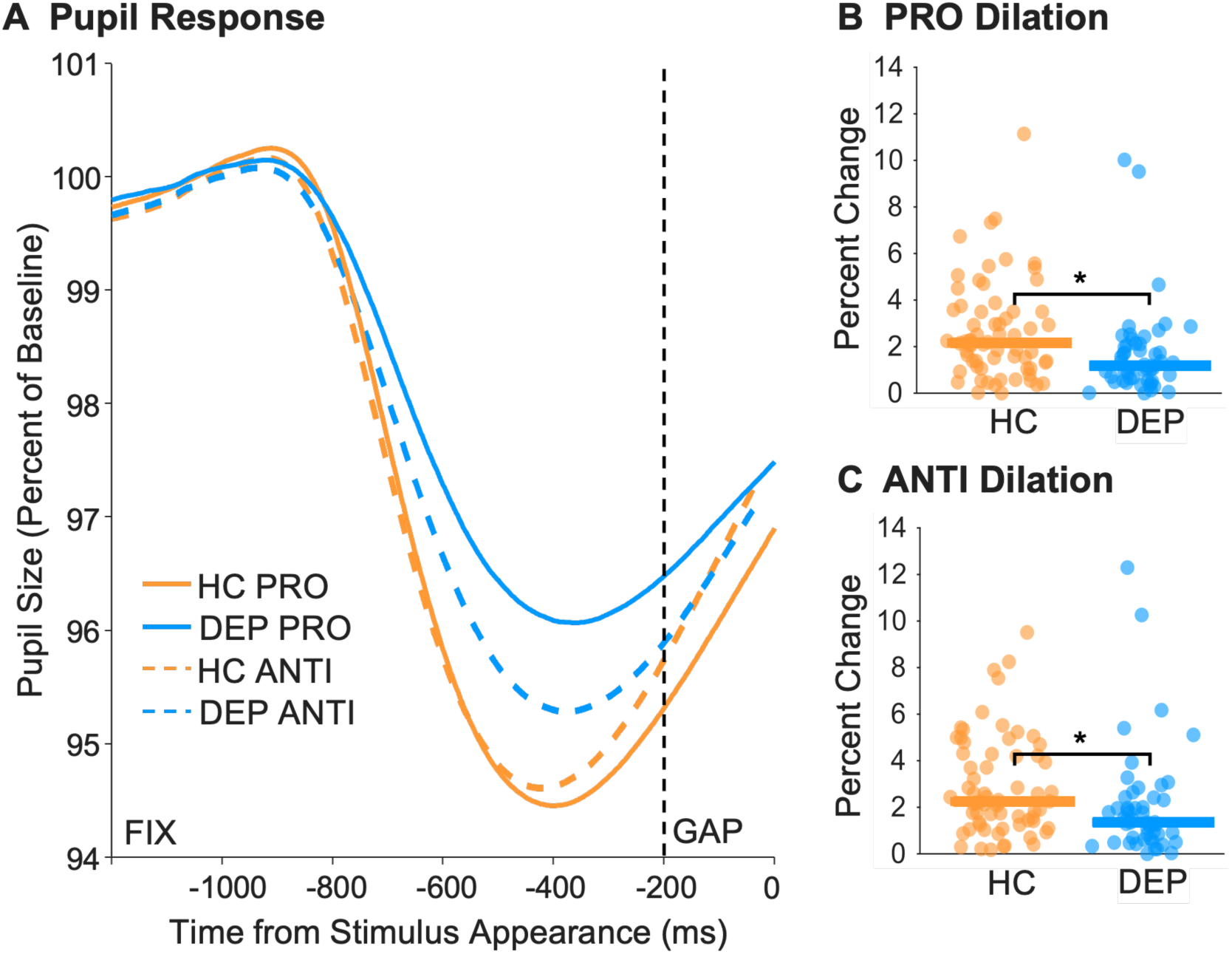
A. Pupil response for pro- and anti-saccade trials following appearance of the fixation point. Vertical dashed line indicates fixation offset. While dilation continues after stimulus appearance, we stop dilation analysis here to avoid saccade-induced artefacts. **B,** C. Individual subject data for dilation size during pro- and anti-saccade trials, respectively. Horizontal lines represents the median group value. *Significant at p<.05 level.

### Associations with Depression Severity and Concurrent Mental Health Symptoms

#### DEP Group

Though it was expected that higher scores on the clinical assessments would correlate with increased oculomotor impairment, most of the significant correlations were in the opposite direction. Within the DEP group, a lower rate of fixation breaks during anti-saccade trials was associated with higher scores on the BSL (*r*=-.367, *p*=.008), GAD-7 (*r*=-.500, *p*<.001), and PHQ-9 (*r*=-.307, *p*=.028). Higher GAD-7 scores also correlated with a lower rate of fixation breaks during pro-saccade trials (*r*=-.431, *p*=002). Higher EDE-Q scores were associated with a higher anti-saccade SRT (*r*=.302, *p*=.031; see Supplementary Table 3).

#### Entire Sample

When analyzing the entire sample, over 30 significant correlations were found between clinical assessments and oculomotor outcomes. Higher SBQ-R scores were correlated with less fixation acquisition (*r*=-.306, *p*<.001). Higher BIS scores were also correlated with less fixation acquisition (*r*=-.279, *p*=.002) as well as smaller pro-saccade pupil dilation size (*r*=-.300, *p*=.003). Higher DERS scores correlated with smaller pro-saccade pupil dilation size (*r*=-.295, *p*=.001) as well as an increased rate of express-latency pro-saccades (*r*=.282, *p*=.002; see Supplementary Table 4).

### Effects of Medication

The majority (88.2%) of the DEP group were on psychotropic medication (antidepressants [38/51], antipsychotics [16/51], and/or stimulants [29/51]). 34/51 were taking more than one class of medication (see Supplementary Fig. 2). Within the DEP group, we did not find any differences on fixation, saccade, or pupil behaviour between those on versus off antidepressants or antipsychotics. No differences were found for fixation or pupil behaviour between those prescribed stimulants, however, those taking stimulants made significantly more express-latency saccades (*F*(1,115)=4.988, *p*=.031, *η²p*=.102) than those who were not. No other effects of stimulants were found for saccade behaviour (see Supplementary – Medication Effects).

## DISCUSSION

The aim of this study was to determine if oculomotor behaviour during a structured video-based eye-tracking task could distinguish adolescents with depression from healthy adolescent controls. We identified several group differences in saccade, fixation, and pupil behaviour during the task. The DEP group 1) made more express-latency saccades overall, 2) had a higher anti-saccade error rate, 3) had a higher rate of fixation breaks, 4) had a delay in fixation acquisition, and 5) had a blunted pupil response. This work extends previous knowledge from adult studies and provides novel insight into how depression affects pupil and fixation behaviour during a pro- and/or anti-saccade task.

When completing IPAST, different neural processes are required for correctly executing pro- versus anti-saccades. Pro-saccades simply rely on visual information to guide the motor behaviour. As expected, there was no group difference in percentage of correct pro-saccade trials, which is consistent with adult findings.^22,24^ Anti-saccades, however, require at least two processes^14^. First, there must be a preemptive, global inhibition of saccades prior to the stimulus appearance to prevent an automatic pro-saccade. Then, participants must generate a voluntary anti-saccade in the opposite direction of the stimulus. This requires removal of the global inhibition, and the voluntary anti-saccade command on one side of the brain must outcompete the automatic pro-saccade command on the opposite side. Proper execution requires cognitive control, with input from the frontal cortex and basal ganglia to the superior colliculus being critical to inhibit the pro-saccade and then generate the voluntary anti-saccade.^47,48^ Due to the additional neural processing required, correct anti-saccades have longer reaction times than correct pro-saccades.^13^

The DEP group had difficulty in top-down global inhibition of automatic pro-saccades, which resulted in more errors on the anti-saccade task (Fig. 3). Their higher error rate was largely driven by an overall increase of express-latency saccades, which are detrimental to anti-saccade performance. Previous studies have found that lesions in areas of the frontal cortex, including the FEF and dlPFC, led to an increase in express-latency saccades and anti-saccade error rates ^17,49^, highlighting its important contributions to correct anti-saccade performance. Given the known association between depression and impairment in the frontal cortex^15^, an increase in express-latency saccades is not unexpected. General difficulty with global inhibition can also explain the increase in fixation breaks in the DEP group. To focus on the fixation cue – i.e., the trial instruction – saccades made to elsewhere or gaze off the screen must be inhibited. Difficulties in maintaining fixation has been found previously in adults with depression, who had reduced fixation stability compared to controls when instructed to maintain gaze on a central stimulus (with and without peripheral distractors).^26,30^ Altogether, these results are indicative of the functional impairments often experienced in depression, in this case difficulties with task focus and cognitive performance.

The second step of making an anti-saccade – generating a voluntary saccade – was not impaired in adolescents with depression. Though depression can be accompanied by motor disturbances, this typically involves slowness or lessened coordination of movement, not prevention.^50^ Indeed, the DEP group could produce correct anti-saccades, as evidenced by their higher than chance correct rate. The lack of group difference in regular-latency anti-saccade errors (Fig. 3C) also demonstrates that they can successfully generate a voluntary saccade in the opposite direction of the stimulus. The ability to generate a voluntary saccade in the absence of a visual stimulus can also be assessed by fixation acquisition behaviour. Although not explicitly instructed, during the ITI (blank screen) participants usually made a saccade to the center of the screen in anticipation of the fixation cue. The ability to do this was not impaired, though the DEP group did so significantly less often than controls (Fig. 2A). As advanced fixation on the center of the screen indicates readiness to receive the trial information, this group difference could be understood as a lack of attention, interest and/or motivation in the DEP group.

Interestingly, we found no group difference in overall SRT. Previous SRT findings are inconsistent in the adult depression literature, but this may be explained by differences in eye-tracking technology (i.e., video-based versus electrooculography) and trial methodology. Importantly, our paradigm included a 200ms gap in between fixation offset and stimulus appearance, which is known to elicit faster SRTs and more express-latency saccades.^51,52^ Prior video-based eye-tracking studies that found a longer SRTs in individuals with depression did not utilize a gap period.^23,28^ Given the DEP group’s tendency to make more express-latency saccades in both pro- and anti-saccade trials, it would be possible that could have counteracted a slower overall SRT. Nonetheless, other studies which did not utilize a gap period found no group differences in SRT.^22,27,30^

Pupil size is adaptive, driven by changes in luminance and arousal, as well as preparatory responses and cognitive control.^45^ As discussed previously, the typical pupil response during IPAST consists of an initial constriction (largely driven by a luminance increase on the fovea) after fixation point appearance, followed by a dilation (driven by saccade preparation and cognitive control) that begins prior to the gap and continues after appearance of the peripheral stimulus. Both trial types require mental preparation, but a larger dilation response is found in preparation for anti-saccade trials in comparison to pro-saccade trials, due to the increase in cognitive control required.^46^ In this study, while we found differences in both dilation and constriction responses, we were most interested in dilation due to its relation to cognitive control. Pupil dilation is coordinated by the superior colliculus, which integrates input from several cortical and subcortical areas, including the frontal cortex, from which preparatory processes and cognitive control signals are sent.^19,46,53^ Since pro-saccade trials were also affected, the difference in dilation likely reflects a deficit in preparatory processing, in addition to the inhibitory control deficits seen on anti-saccade trials alone. Similarly, altered dilation responses in depression have been found in other eye-tracking tasks. This includes a reduced dilation response in anticipation of reward^54^, and a larger dilation response when viewing negative stimuli.^55^

Though several correlations between clinical variables and oculomotor behaviour were identified within the DEP group (see Supplementary Table 3), we highlight a couple which we deem particularly relevant. Surprisingly, the only correlation between depression severity (PHQ) and oculomotor behaviour was for anti-saccade fixation breaks. A higher PHQ score correlated with less fixation breaks, which is the opposite direction than expected, especially given the identified group differences. It is possible that the more severely depressed adolescents did not accurately rate their depression level in the PHQ, due to difficulty recognizing a departure from “normal” mood and behaviour or difficulties with identifying/understanding their emotions (e.g., alexithymia; a common occurrence in depression)^56^. This may be further substantiated by the documented discrepancies in self-rated depression severity and clinician ratings.^57^ Another explanation is that in the more severe cases, medication could be alleviating mood and behavioural symptoms, but not cognitive impairments. This issue has been reported previously^58,59^, and could explain why those with lower PHQ scores are still experiencing difficulty with task focus. Additionally, anxiety severity (GAD) was negatively correlated with rate of fixation breaks in both pro- and anti-saccade trials. This result was initially surprising, as we would expect an increase in clinical severity to correlate with a decrease in performance. However, previous research has found that a moderate to high levels of arousal, relative to low levels, can improve task performance depending on task difficulty.^60^ Thus, anxiety scores may have acted as a protective factor in this case, with individuals who are more anxious being more cautious and focused on the fixation point instruction.

## Limitations

There are some important limitations to this study. Because adolescents with depression were recruited from a hospital outpatient site, results may not be generalizable to milder cases or non-treatment seeking individuals. Additionally, to have a clinically representative sample, we included participants with psychiatric comorbidities and those taking medications. This approach allowed us to examine associations between eye-tracking measures and concurrent symptoms and thus capture the complexity of clinical “real world” presentations. However, the results may not be generalizable to populations without comorbidities or to individuals not receiving pharmacological treatment. Finally, the clinical group had a relatively high proportion of gender-diverse individuals (20%). Despite our ability to closely match our control group based on age and sex assigned at birth, we had a much lower proportion of gender diverse adolescents in our control group (3%). The effect of gender identity – as opposed to sex – on oculomotor behaviour has not yet been studied to our knowledge and necessitates detailed investigation.

There remains substantial variability in the methodology within the eye-tracking field, and even so within pro- and anti-saccade paradigms. Previous studies have used different eye-tracking techniques, trial structures, and task structures – some having separate blocks for pro-versus anti-saccade trials, some, like ours, having interleaved trials, and some not having pro- or anti-saccade trials at all. Our approach is strengthened by a large number of trials (120 pro-saccade and 120-anti-saccade) and relatively large clinical sample size (51 DEP) for the field, with most of the aforementioned studies employing smaller numbers of participants ^21–25,27–29^ and/or trials.^21–24,26,29,30^ While these studies provided key information, our increased numbers may allow for more statistical power. Additionally, randomly interleaving pro- and anti-saccade trials rather than having pure pro- and anti-saccade blocks allow us to better investigate inhibitory control. Interleaved trials require constant task-switching as you are unaware what the next trial will be, in comparison to a predictable anti-saccade block where it is learned to always look away from the stimulus. Finally, in addition to novel fixation and pupil analysis, we further divided anti-saccade errors into express- and regular-latency errors.^13^ This separation is rarely done in the field, but this distinction is important given the different neural processes behind these behaviours. In this case, the distinction was particularly significant given we found a group difference in express- but not regular-latency errors, which gives further insight into the depression group’s behaviour.

## CONCLUSION

The present study identified several differences in oculomotor behaviour between adolescents with depression and healthy controls during an interleaved pro- and anti-saccade task. In addition to an increase in anti-saccade errors and express-latency saccades in pro- and anti-saccade trials, we identified difficulties with acquiring and maintaining fixation as well as an altered pupillary response during the trial instruction epoch. These findings may bring new insight into the oculomotor phenotype of depression and highlight the potential of including fixation and pupil behaviour in clinical eye movement research. Taken together, these oculomotor alterations could be utilized in the future as a tool for early identification in at-risk youth.

## Data Availability

Data generated or analyzed during this study are available from the corresponding author upon reasonable request.

