## Supplementary Material for "Identifying an oculomotor phenotype for adolescent depression with an interleaved pro- and anti-saccade task"

### Supplementary Methods

In the Interleaved Pro- and Anti-Saccade Task (IPAST), we measure pupil behaviour during the FIX and GAP epochs, where we see an initial constriction quickly followed by a dilation. This pattern of response (see pink curve below) occurs across all ages, and across different clinical populations[1-5]. However, the magnitude of constriction and/or dilation varies. This is because constriction and dilation responses, which are influenced by several factors (e.g., luminance, arousal, cognitive control), are driven by brain areas which develop/change with age and are impacted by neurodegenerative and psychiatric disorders[6].

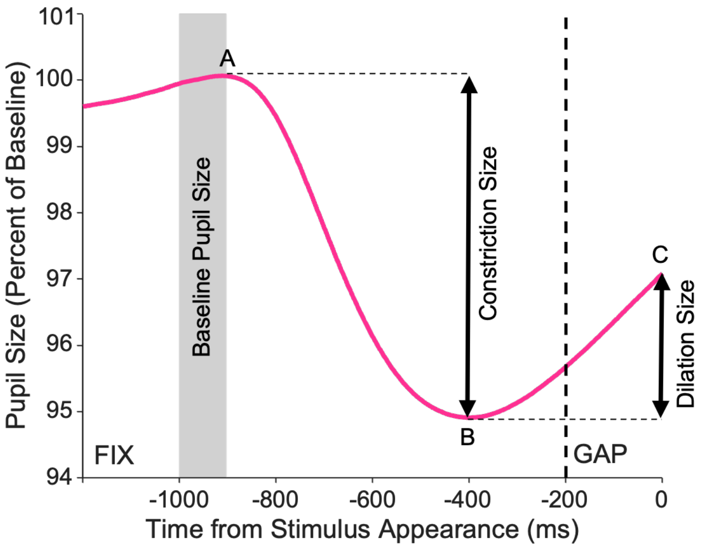

**Supplementary Figure 1.** Example of a typical pupillary response curve during IPAST. Baseline pupil size is calculated as the average size within the grey bar. Constriction and dilation measurements are indicated by the two arrows. Constriction is measured as the difference between B and A, and dilation is measured as the difference between C and B. Data is represented in percent change as opposed to pixels to correct for differences in baseline pupil size.

**Supplementary Results - IPAST Behaviour**

As discussed in the main text, the depression group displayed altered fixation, saccade, and pupil behaviour during the task. These included difficulties acquiring and maintaining fixation, an increased express-latency saccade rate for pro- and anti-saccade trials, an increase in anti-saccade errors, a smaller constriction response for pro-saccade trials, and a smaller dilation response for pro- and anti-saccade trials. Group means, standard deviations, and statistics for all oculomotor outcomes mentioned in main text can be found below.

**Supplementary Table 1.** Between-group analysis of fixation, saccade, and pupil behaviour.

| | **HC (n=66)** | **DEP (n=51)** | | | | |
|---|---|---|---|---|---|---|
| | *M (SD)* | *M (SD)* | *F* | *df* | *p* | $\eta^2 p$ |
| **Fixation Behaviour** | | | | | | |
| PRO Fixation Breaks (%) | 5.50 (6.10) | 8.49 (7.93) | 5.341 | 1, 115 | **.023*** | .044 |
| ANTI Fixation Breaks (%) | 5.68 (7.26) | 10.46 (10.57) | 8.384 | 1, 115 | **.005**** | .068 |
| Fixation Acquisition (%) | 86.39 (9.68) | 77.52 (14.16) | 16.136 | 1, 115 | **<.001**** | .123 |
| **Saccade Behaviour** | | | | | | |
| PRO SRT (ms) | 170.00 (31.88) | 159.27 (31.82) | 3.263 | 1, 115 | .073 | .028 |
| ANTI SRT (ms) | 244.15 (37.44) | 245.53 (41.32) | 0.036 | 1, 115 | .851 | .000 |
| PRO Correct Rate (%) | 97.72 (2.45) | 97.52 (2.82) | 0.169 | 1, 115 | .682 | .001 |
| ANTI Correct Rate (%) | 76.93 (14.92) | 69.43 (17.12) | 6.378 | 1, 115 | **.013*** | .053 |
| PRO Express Correct (%) | 37.31 (23.57) | 48.65 (25.04) | 6.310 | 1, 115 | **.016*** | .052 |
| ANTI Express Errors (%) | 13.28 (11.99) | 19.79 (16.90) | 5.930 | 1, 115 | **.016*** | .049 |
| ANTI Regular Errors (%) | 9.79 (7.65) | 10.77 (9.28) | 0.397 | 1, 115 | .530 | .003 |
| **Pupil Behaviour** | | | | | | |
| Baseline Pupil Size (px) | 4004.18 (1325.59) | 4038.08 (1301.77) | 0.19 | 1, 115 | .890 | .000 |
| PRO Constriction Size (% change) | -6.27 (4.77) | -4.36 (5.50) | 4.039 | 1, 115 | **.047*** | .034 |
| PRO Dilation Size (% change) | 2.65 (2.10) | 1.67 (1.91) | 6.768 | 1, 115 | **.011*** | .056 |
| ANTI Constriction Size (% change) | -6.06 (4.64) | -5.04 (6.68) | .944 | 1, 115 | .333 | .008 |
| ANTI Dilation Size (% change) | 2.88 (2.10) | 2.04 (2.32) | 4.251 | 1, 115 | **.041*** | .036 |

HC=healthy control, DEP=depression group, PRO=pro-saccade trials, ANTI=anti-saccade trials. SRT=Saccadic reaction time. *Significant at the $p<.05$ level. ** Significant at the $p<.01$ level.

**Supplementary Results - Correlations**

The supplementary table below displays correlations between oculomotor behaviour and clinical scores within the depression group. Curiously, apart from the correlation between EDEQ scores and anti-saccade reaction time, these significant correlations are in the opposite direction than expected (i.e., greater clinical impairment correlated with less oculomotor impairment). Some potential explanations behind these surprising results are suggested in the main text discussion.

**Supplementary Table 2.** Correlations between oculomotor behaviour and clinical scores for the depression group (n=51).

| | **BIS (n=51)** | **BSL (n=51)** | **DERS (n=51)** | **EDE-Q (n=51)** | **GAD (n=51)** | **PHQ (n=51)** | **SBQ-R (n=49)** |
|---|---|---|---|---|---|---|---|
| **Fixation Behaviour** | | | | | | | |
| PRO Fixation Breaks (%) | *r*=.010<br>*p*=.942 | *r*=-.236<br>*p*=.096 | *r*=-.128<br>*p*=.371 | *r*=-.030<br>*p*=.835 | *r*=-.431<br>***p*=.002** | *r*=-.142<br>*p*=.320 | *r*=.102<br>*p*=.413 |
| ANTI Fixation Breaks (%) | *r*=-.069<br>*p*=.632 | *r*=-.367<br>***p*=.008** | *r*=-.132<br>*p*=.357 | *r*=-.078<br>*p*=.586 | *r*=-.500<br>***p*<.001** | *r*=-.307<br>***p*=.028** | *r*=.127<br>*p*=.384 |
| Fixation Acquisition (%) | *r*=-.151<br>*p*=.291 | *r*=.133<br>*p*=.353 | *r*=.219<br>*p*=.123 | *r*=.038<br>*p*=.793 | *r*=.137<br>*p*=.339 | *r*=.240<br>*p*=.090 | *r*=-.096<br>*p*=.510 |
| **Saccade Behaviour** | | | | | | | |
| PRO SRT (ms) | r=-.059<br>p=.678 | *r*=-.117<br>*p*=.412 | *r*=-.092<br>*p*=.520 | *r*=.022<br>*p*=.880 | *r*=.092<br>*p*=.522 | *r*=-.180<br>*p*=.207 | *r*=.006<br>*p*=.965 |
| ANTI SRT (ms) | r=.124<br>p=.386 | *r*=.044<br>*p*=.760 | *r*=.037<br>*p*=.798 | *r*=.302<br>***p*=.031*** | *r*=.097<br>*p*=.500 | *r*=-.046<br>*p*=.751 | *r*=.127<br>*p*=.386 |
| PRO Correct Rate (%) | r=-.004<br>p=.975 | *r*=-.066<br>*p*=.644 | *r*=.048<br>*p*=.739 | *r*=.003<br>*p*=.985 | *r*=-.180<br>*p*=.207 | *r*=.096<br>*p*=.503 | *r*=-.134<br>*p*=.358 |
| ANTI Correct Rate (%) | r=-.215<br>p=.130 | *r*=-.066<br>*p*=.646 | *r*=.003<br>*p*=.981 | *r*=-.011<br>*p*=.941 | *r*=.090<br>*p*=.529 | *r*=.029<br>*p*=.842 | *r*=-.102<br>*p*=.484 |
| PRO Express Correct (%) | r=.150<br>p=.293 | *r*=.160<br>*p*=.261 | *r*=.163<br>*p*=.254 | *r*=.047<br>*p*=.746 | *r*=-.033<br>*p*=.819 | *r*=.138<br>*p*=.334 | *r*=.149<br>*p*=.308 |
| ANTI Express Errors (%) | r=.201<br>p=.157 | *r*=.124<br>*p*=.387 | *r*=.073<br>*p*=.611 | *r*=.003<br>*p*=.985 | *r*=-.062<br>*p*=.667 | *r*=.047<br>*p*=.742 | *r*=.077<br>*p*=.600 |
| ANTI Regular Errors (%) | r=.030<br>p=.833 | *r*=-.104<br>*p*=.469 | *r*=-.139<br>*p*=.329 | *r*=.015<br>*p*=.918 | *r*=-.054<br>*p*=.706 | *r*=-.139<br>*p*=.332 | *r*=.049<br>*p*=.737 |
| **Pupil Behaviour** | | | | | | | |
| Baseline Pupil Size (px) | *r*=-.161<br>*p*=.259 | *r*=.001<br>*p*=.997 | *r*=-.075<br>*p*=.600 | *r*=-.133<br>*p*=.353 | *r*=-.028<br>*p*=.843 | *r*=-.070<br>*p*=.624 | *r*=-.069<br>*p*=.637 |
| PRO Constriction Size (% change) | *r*=-.007<br>*p*=.960 | *r*=.102<br>*p*=.477 | *r*=.117<br>*p*=.414 | *r*=-.081<br>*p*=.574 | *r*=-.052<br>*p*=.718 | *r*=.051<br>*p*=.720 | *r*=.121<br>*p*=.408 |
| PRO Dilation Size (% change) | *r*=-.079<br>*p*=.581 | *r*=-.128<br>*p*=.372 | *r*=-.168<br>*p*=.239 | *r*=.008<br>*p*=.954 | *r*=-.041<br>*p*=.776 | *r*=-.091<br>*p*=.526 | *r*=-.083<br>*p*=.568 |
| ANTI Constriction Size (% change) | *r*=-.045<br>*p*=.756 | *r*=.102<br>*p*=.478 | *r*=-.097<br>*p*=.496 | *r*=-.101<br>*p*=.480 | *r*=-.039<br>*p*=.785 | *r*=.069<br>*p*=.633 | *r*=.042<br>*p*=.773 |
| ANTI Dilation Size (% change) | *r*=-.033<br>*p*=.820 | *r*=-.093<br>*p*=.517 | *r*=-.092<br>*p*=.521 | *r*=.080<br>*p*=.579 | *r*=.010<br>*p*=.946 | *r*=-.019<br>*p*=.896 | *r*=-.097<br>*p*=.506 |

BIS=Barratt Impulsiveness Scale, BSL=Borderline Symptom List, DERS=Difficulties in Emotion Regulation Scale, EDE-Q=Eating Disorder Examination Questionnaire, GAD=Generalized Anxiety Disorder Scale, PHQ=Patient Health Questionnaire, SBQ-R=Suicide Behaviors Questionnaire Revised. *Significant at the p<.05 level. **Significant at the p<.01 level.

The supplementary table below displays correlations between oculomotor behaviours and clinical scores for the combined healthy control and depression groups. When adding the control group, many more significant correlations emerged. We believe this is largely driven by the large group differences in clinical scores.

**Supplementary Table 3.** Correlations between oculomotor behaviour and clinical scores for the entire sample (N=117).

| | **BIS (N=117)** | **BSL (N=117)** | **DERS (N=117)** | **EDE-Q (N=117)** | **GAD (N=117)** | **PHQ (N=117)** | **SBQ-R (N=115)** |
|---|---|---|---|---|---|---|---|
| **Fixation Behaviour** | | | | | | | |
| Fixation Breaks – PRO (%) | r=.219 **p=.018*** | r=.060 p=.523 | r=.145 p=.118 | r=.142 p=.127 | r=.027 p=.771 | r=.142 p=.126 | r=.198 **p=.036*** |
| Fixation Breaks – ANTI (#) | r=.187 p=.044 | r=.036 p=.698 | r=.164 p=.078 | r=.154 p=.097 | r=.036 p=.704 | r=.112 p=.229 | r=.237 **p=.011*** |
| Fixation Acquisition (%) | r=-.279 **p=.002**** | r=-.201 **p=.030*** | r=-.178 p=.055 | r=-.067 p=.475 | r=-.222 **p=.016*** | r=-.165 p=.075 | r=-.306 **p<.001**** |
| **Pupil Behaviour** | | | | | | | |
| Baseline Pupil Size (px) | r=.093 p=.318 | r=.002 p=.987 | r=.034 p=.715 | r=-.116 p=.211 | r=.022 p=.812 | r=.012 p=.896 | r=-.026 p=.779 |
| Constriction Size – PRO (% change) | r=.276 **p=.003**** | r=.196 p=.034 | r=.248 **p=.007**** | r=-.006 p=.950 | r=.149 p=.108 | r=.179 p=.053 | r=.145 p=.121 |
| Dilation Size – PRO (% change) | r=-.300 **p=.001**** | r=-.239 **p=.009**** | r=-.295 **p=.001**** | r=-.110 p=.238 | r=-.249 **p=.007**** | r=-.236 **p=.011*** | r=-.205 **p=.028*** |
| Constriction Size – ANTI (% change) | r=.145 p=.120 | r=.123 p=.188 | r=.183 p=.049 | r=-.039 p=.678 | r=.075 p=.422 | r=.111 p=.233 | r=.044 p=.641 |
| Dilation Size – ANTI (% change) | r=-.248 **p=.007**** | r=-.197 **p=.033*** | r=-.222 **p=.016*** | r=-.056 p=.546 | r=-.187 **p=.044*** | r=-.181 p=.051 | r=-.186 **p=.046*** |
| **Saccade Behaviour** | | | | | | | |
| SRT – PRO (ms) | r=-.119 p=.200 | r=-.198 **p=.038*** | r=-.222 **p=.016*** | r=-.105 p=.259 | r=-.144 p=.123 | r=-.236 **p=.011*** | r=-.095 p=.311 |
| SRT – ANTI (ms) | r=.061 p=.511 | r=.010 p=.915 | r=.001 p=.993 | r=.103 p=.270 | r=.025 p=.788 | r=-.030 p=.748 | r=.076 p=.420 |
| Correct Rate – PRO (%) | r=.018 p=.851 | r=-.071 p=.445 | r=-.019 p=.836 | r=-.038 p=.685 | r=-.084 p=.368 | r=.003 p=.975 | r=-.109 p=.246 |
| Correct Rate – ANTI (%) | r=-.191 **p=.039*** | r=-.219 **p=.018*** | r=-.234 **p=.011*** | r=-.076 p=.413 | r=-.198 **p=.033*** | r=-.189 **p=.041*** | r=-.209 **p=.025*** |
| Correct Express – PRO (%) | r=.114 p=.122 | r=.266 **p=.004**** | r=.282 **p=.002**** | r=.143 p=.124 | r=.206 **p=.026*** | r=.268 **p=.003**** | r=.225 **p=.016*** |
| Express Errors – ANTI (%) | r=.159 p=.088 | r=.248 **p=.007**** | r=.236 **p=.010**** | r=.100 p=.283 | r=.200 **p=.031*** | r=.214 **p=.020*** | r=.197 **p=.035*** |
| Regular Errors – ANTI (%) | r=.094 p=.313 | r=.007 p=.941 | r=.043 p=.643 | r=-.027 p=.776 | r=.035 p=.705 | r=-.007 p=.942 | r=.063 p=.503 |

*Significant at the p<.05 level. **Significant at the p<.01 level.

### Supplementary Results - Medication Effects

Due to reported effects of psychotropic medications on oculomotor behaviour, particularly pupil dilation[7], we wanted to investigate if there were within-group differences between patients on versus off antidepressants, antipsychotics, and stimulants. While we were also interested in looking at how interactions of these medications affected oculomotor behaviour, the sample sizes for most of these interactions were too low to meaningfully investigate. The Venn diagram to the right (Supp. Fig. 2) displays the medication statuses of the participants in the depression group (n=51).

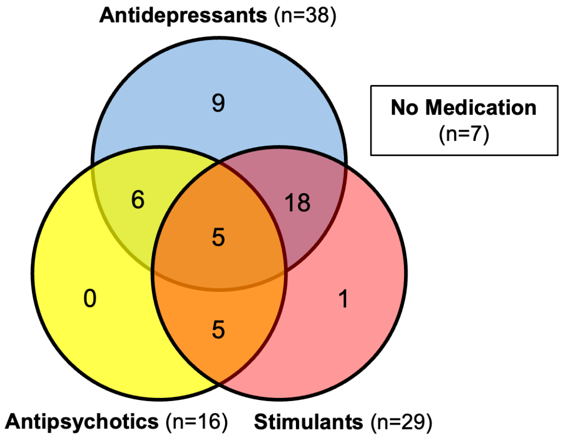

**Supplementary Figure 2.** Number of patients taking each class of medication.

Previous studies in our laboratory employed the ON versus OFF method when investigating effects of psychotropic medication on IPAST behaviour in youth with eating disorders[8] and a predictive saccade task in borderline personality disorder (BPD)[9]. In eating disorders, it was found that medication usage (antidepressants, antipsychotics, benzodiazepines) correlated with increased baseline pupil size but did not identify any medication effects on saccade or blink behaviour. In BPD, no effects of antidepressants, antipsychotics, or stimulants on pupil size or blink behaviour were identified.

We ran three separate MANOVAs for fixation (Supp. Table 4), saccade (Supp. Table 5), and pupil (Supp. Table 6) outcomes with the medication classes (antidepressants, antipsychotics, and stimulants) as fixed factors. Medication status for each class was coded as ON or OFF. No differences were identified for any fixation or pupil outcomes between those ON/OFF antidepressants, antipsychotics, or stimulants. Across saccade outcomes, we found one effect of medication status. Patients taking stimulants made significantly more correct express saccades (55.4%) than those not taking stimulants (39.8%; $F$=4.988(1,49), $p$=.031, $\eta^2 p$=.102). However, given that no other oculomotor outcomes were affected by stimulant status, we do not believe that they are driving the overall deficit in cognitive control which we see across fixation, saccade, and pupil behaviour in the depression group.

**Supplementary Table 4.** Within group-differences in fixation behaviour between patients on versus off medication.

| | ON | OFF | | | | |
|---|---|---|---|---|---|---|
| | *M (SD)* | *M (SD)* | *F* | *df* | *p* | $\eta^2p$ |
| **Antidepressants (38 ON, 13 OFF)** | | | | | | |
| PRO Fixation Breaks (%) | 8.23 (7.78) | 9.27 (8.62) | 0.25 | 1, 49 | .874 | .001 |
| ANTI Fixation Breaks (%) | 9.30 (7.91) | 13.86 (15.98) | .436 | 1, 49 | .513 | .010 |
| Fixation Acquisition (%) | 78.04 (14.07) | 76.02 (14.89) | .339 | 1, 49 | .564 | .008 |
| **Antipsychotics (16 ON, 35 OFF)** | | | | | | |
| PRO Fixation Breaks (%) | 7.72 (8.05) | 8.84 (7.96) | .220 | 1, 49 | .641 | .005 |
| ANTI Fixation Breaks (%) | 7.76 (6.59) | 11.69 (11.83) | .009 | 1, 49 | .924 | .000 |
| Fixation Acquisition (%) | 79.20 (14.42) | 76.76 (14.18) | .185 | 1, 49 | .669 | .004 |
| **Stimulants (29 ON, 22 OFF)** | | | | | | |
| PRO Fixation Breaks (%) | 7.72 (6.21) | 9.52 (9.81) | 1.140 | 1, 49 | .291 | .025 |
| ANTI Fixation Breaks (%) | 9.96 (7.78) | 11.11 (13.57) | .724 | 1, 49 | .399 | .016 |
| Fixation Acquisition (%) | 74.24 (14.80) | 81.86 (12.27) | 1.068 | 1, 49 | .307 | .024 |

ON=patients taking the medication, OFF=patients not taking the medication.

**Supplementary Table 5.** Within-group differences in saccade behaviour between patients on versus off medication.

| | ON | OFF | | | | |
|---|---|---|---|---|---|---|
| | *M (SD)* | *M (SD)* | *F* | *df* | *p* | $\eta^2p$ |
| **Antidepressants (38 ON, 13 OFF)** | | | | | | |
| PRO SRT (ms) | 159.58 (34.61) | 158.36 (22.91) | .000 | 1, 49 | .995 | .000 |
| ANTI SRT (ms) | 243.63 (42.21) | 251.08 (39.72) | .021 | 1, 49 | .885 | .000 |
| PRO Correct Rate (%) | 97.15 (3.09) | 98.62 (1.41) | 3.458 | 1, 49 | .070 | .073 |
| ANTI Correct Rate (%) | 68.88 (19.16) | 71.04 (9.25) | .395 | 1, 49 | .533 | .009 |
| PRO Express Correct (%) | 49.54 (26.70) | 46.05 (20.08) | .149 | 1, 49 | .702 | .003 |
| ANTI Express Errors (%) | 20.83 (18.49) | 16.75 (11.10) | .681 | 1, 49 | .702 | .003 |
| ANTI Regular Errors (%) | 10.28 (9.64) | 12.21 (8.32) | .121 | 1, 49 | .729 | .003 |
| **Antipsychotics (16 ON, 35 OFF)** | | | | | | |
| PRO SRT (ms) | 162.98 (40.83) | 157.58 (27.28) | 1.229 | 1, 49 | .274 | .027 |
| ANTI SRT (ms) | 252.47 (44.38) | 242.36 (40.11) | .801 | 1, 49 | .376 | .018 |
| PRO Correct Rate (%) | 97.03 (3.57) | 97.75 (2.43) | .465 | 1, 49 | .499 | .010 |
| ANTI Correct Rate (%) | 72.09 (17.00) | 68.22 (17.29) | .208 | 1, 49 | .651 | .005 |
| PRO Express Correct (%) | 49.99 (28.38) | 48.04 (23.77) | .562 | 1, 49 | .457 | .013 |
| ANTI Express Errors (%) | 19.09 (19.05) | 20.11 (16.12) | .374 | 1, 49 | .544 | .008 |
| ANTI Regular Errors (%) | 8.82 (6.90) | 11.67 (10.14) | .076 | 1, 49 | .784 | .002 |
| **Stimulants (29 ON, 22 OFF)** | | | | | | |
| PRO SRT (ms) | 152.74 (29.37) | 167.88 (33.53) | 3.289 | 1, 49 | .077 | .070 |
| ANTI SRT (ms) | 242.73 (41.71) | 249.23 (41.49) | .375 | 1, 49 | .544 | .008 |
| PRO Correct Rate (%) | 97.87 (2.22) | 97.06 (3.46) | .492 | 1, 49 | .487 | .011 |
| ANTI Correct Rate (%) | 65.23 (19.44) | 74.98 (11.72) | 1.829 | 1, 49 | .183 | .040 |
| PRO Express Correct (%) | 55.35 (25.12) | 39.81 (22.51) | 4.988 | 1, 49 | **.031*** | .102 |
| ANTI Express Errors (%) | 24.10 (18.57) | 14.11 (12.70) | 3.260 | 1, 49 | .078 | .069 |
| ANTI Regular Errors (%) | 10.67 (10.74) | 10.91 (7.14) | .643 | 1, 49 | .427 | .014 |

*Significant at the p<.05 level.

**Supplementary Table 6.** Within-group differences in pupil behaviour between patients on versus off medication.

| | ON | OFF | | | | |
|---|---|---|---|---|---|---|
| | *M (SD)* | *M (SD)* | *F* | *df* | *p* | $\eta^2 p$ |
| **Antidepressants (38 ON, 13 OFF)** | | | | | | |
| Baseline Pupil Size (px) | 4185.93 (1351.33) | 3605.90 (1076.52) | .555 | 1, 49 | .460 | .012 |
| PRO Constriction Size (% change) | -4.37 (6.03) | -4.35 (3.76) | .003 | 1, 49 | .958 | .000 |
| PRO Dilation Size (% change) | 1.86 (2.13) | 1.11 (0.84) | .429 | 1, 49 | .516 | .010 |
| ANTI Constriction Size (% change) | -5.20 (7.41) | -4.57 (3.98) | .036 | 1, 49 | .850 | .001 |
| ANTI Dilation Size (% change) | 2.15 (2.59) | 1.71 (1.23) | .029 | 1, 49 | .865 | .001 |
| **Antipsychotics (16 ON, 35 OFF)** | | | | | | |
| Baseline Pupil Size (px) | 4435.65 (1350.59) | 3856.33 (1256.56) | 1.189 | 1, 49 | .281 | .026 |
| PRO Constriction Size (% change) | -3.95 (4.22) | -4.55 (6.04) | .032 | 1, 49 | .858 | .001 |
| PRO Dilation Size (% change) | 1.19 (0.89) | 1.89 (2.20) | 1.436 | 1, 49 | .237 | .032 |
| ANTI Constriction Size (% change) | -4.90 (4.91) | -5.11 (7.41) | .133 | 1, 49 | .717 | .003 |
| ANTI Dilation Size (% change) | 1.36 (0.83) | 2.35 (2.70) | 1.167 | 1, 49 | .286 | .026 |
| **Stimulants (29 ON, 22 OFF)** | | | | | | |
| Baseline Pupil Size (px) | 4096.75 (1226.16) | 3960.74 (1420.96) | .060 | 1, 49 | .808 | .001 |
| PRO Constriction Size (% change) | -4.29 (4.66) | -4.45 (6.56) | .289 | 1, 49 | .593 | .007 |
| PRO Dilation Size (% change) | 1.79 (1.78) | 1.51 (2.09) | .558 | 1, 49 | .459 | .013 |
| ANTI Constriction Size (% change) | -4.83 (5.91) | -5.32 (7.71) | .463 | 1, 49 | .500 | .010 |
| ANTI Dilation Size (% change) | 2.11 (2.09) | 1.94 (2.64) | .142 | 1, 49 | .708 | .003 |
